# Evaluation of the Curitiba University intern medical students’ hability in breaking bad news in a OSCE test

**DOI:** 10.1101/2023.01.20.23284662

**Authors:** Thiago Ribeiro E Silva, João Paulo Zanardini De Lara, Leonardo Ajuz Do Prado Oliveira, Lucas Baggio, Matheus Pisa Freitas, Rodrigo Ribeiro E Silva, João Pedro Ribeiro Baptista, Marcos Takimura, Cristina Terumy Okamoto

## Abstract

**Objective:** Breaking bad news (BBN) has a big influence in patients’ lives, but still a lot of health care providers struggle when doing it with efficiency and empathy.

**Methods:** This study is a prospective cohort that evaluated the knowledge and the evolution of fifth year medical students in breaking bad news, and investigated which factors had a positive or negative influence on their scores. Two simulations were conducted during the year in the model of Objective Structured Clinical Examination (OSCE), the students’ scores were acquired through a checklist based on the SPIKES protocol. Posteriorly, an online questionnaire was filled by the students, containing objective and open questions relevant to the BBN scenario.

**Results:** Participated in this study 85 undergraduates, in the first OSCE 52% (n = 44) scored above the minimum institutional average, in the second OSCE 59% (n = 50) scored above the minimum institutional average. Comparing both activities there were isolated differences between isolated items at the checklist, but without statistical significance. The factor that had a positive influence for scoring higher was having previous experience in BBN, during the second OSCE the group that had experience scored an average of 3,11 points, the other group had an average of 2,57 only (p = 0,012).

**Conclusion:** The performance was median, and no score evolution was observed between the activities. Previous experience was the more important factor for a better score.

**ILUSTRATIONS’ LIST:** Table 1 Number of students’ right scores, per criteria in both of simulations

## INTRODUCTION

The patient-physician relationship is a very important concept. Despite advenced in techonogies, nothing is capable to substitute a clear comunication (VICTORINO et al., 2007). Empathy and comprehension, as well as health professional’ s emotional control are all vital skills to let the patient and her/his relatives to understand diagnosis, healthy stats, and prognosis. All these skills are also important to provide an confortable enviroment, whereas patient’ s emotions are taking seriously. (BORGES; FREITAS; GURGEL, 2012; JUCÁ et al., 2010; VICTORINO et al., 2007).

Bad news are defined as all kind of anoucement – terminal ill or chronic disease, for example - capable to change radicaly or negatively patient’ s view about her/his own desteny, life, and future. (SUCUPIRA, 2007; VICTORINO et al., 2007). The act of ignore or the act of non-comprehension psychologic impacts caused by bad informations can lead to emotional stress and feelings like anxiety, exhaustive, angry, and guilty. (DEAN; WILLIS, 2016).

All this painful process of accept bad news can be facilitate if the physician has experience. The health professional difficulty in breaking bad news is still a problem regonize in the hole world, though. There are a lot of reasons for that, including physician’ s own fears, like fear of develop the patient’ s misery, fear to feel or to caus unconfortable situations, as well as guilty by the relatives and by the patient, and fear to judicial processes. (DEAN; WILLIS, 2016; SOMBRA NETO et al., 2017). Because of those reasons CMN can easily become unconfortble and anti-ethics situations (SOMBRA NETO et al., 2017).

There are some of CMN strategies criated to facilitate this moment to physicians. The most famous one is known as SPIKES protocol. The protocol’ s name is a abreviation of six items criated primarily for using with oncologyc patients, but now a days it has been adapted and allowed to use to breaking all types of bad news to any kind of patient. (BAILE et al., 2000; DEAN; WILLIS, 2016; SEIFART et al., 2014). SPIKES goes for: *Setting* to prepare the place to the patient with time and privacy for this conversationç *Perception* to understand how many things does the patient know about her/his own disease; *Invitation*, to question and respect the amount of information and details that the patient really wants to know; *Knowledge* to communicate the bad new and have the hability to explain all the informations, always in agreement with the other rules. For this particular moment it is reccomended to use introduction sentances that indicate to the patient that bad news are coming; *Emotions* to let the patient to feel her/his feelings. The physician has to be empathic and offer confort, respecting moments of silence; S*trategy and summary* to plan treatments and prognosis, as well as guarantee that the patient understands everything that has been discussed in this meeting (BAILE et al., 2000).

Teaching breaking bad news in medical schools is extremely important. The National Education Comitee (CNE) proposed changes in the medical education of all medical schools in Brazil. It was propose that the schools teach verbal and non-verbal communication, and techniques of breaking bad news. (BRASIL. MINISTÉRIO DA EDUCAÇÃO. CONSELHO NACIONAL DE EDUCAÇÃO, 2014). This subject is known as “Breaking Bad News” and is an obrigatory subject to all medical students in Brazil. Recently new strategies were criated to improve the learning of students. These new methods has each one pros and cons, and there are some examples of these new strategies, as theory classes, simulation using other medical student, patient simulator (PS) and observe real moments happening inside the hospital (NONINO; MAGALHÃES; FALCÃO, 2012; ROSENBAUM; FERGUSON; LOBAS, 2004).

The use of PS for teaching some specifics subjects is growing up fast. One of the most exemples of PS is called OSCE (*Objective Structured Clinical Examinations*) (ROSENBAUM; FERGUSON; LOBAS, 2004). OSCE is an evalution type of method using in medical students that is based in to simulate the reality. It was first described by Harden in 1975 (HARDEN et al., 1975). This method is composed of stations that students has to rotate. Each station have their own goals and lead to the student to show skills of specific clinic situations. This activity is followed by a debriefing, a reuion that medical students receive a feedback about their performance by the doctors whom is observing them the hole time at all the stations. (HARDEN et al., 1975). OSCE is a new resource of learning that allows medical students to train their habilities in a safe space, without expose them to patients and ocasionaly build unconfortable situations (DOURADO; GIANNELLA, 2014; SCHWELLER et al., 2018).

This study was made with medical students in the last two years of medical school and has the following prupose: to evaluate if the basic and clinic phases of med school is preparing their students to breaking bad news and use of SPIKES protocol; to observe evalution of the students in this particular skill in the OSCE evaluation with P; to use checklists as evaluation stool related about SPIKES protocol; to understand how import is this training in medical students life.

## METHODS

This study is a prospective coorte type study and envolved medical students (men and women, an amount of 106 students) in the 5^th^ year of Medical School of Universidade Positivo, located in Curitiba-PR. It was developed in the year of 2019 and it lasted one year. All the studentes signed the informed consent form for both online and in class activities. The study was approved by the Universidade Positivo’ s ethics comitee over the number CAAE 16278619.2.0000.0093 (appendix 3), as well as University’ s dean authorization (appendix 4).

Students whom fullfiled all the steps of this project participated of this study (an amount of 85 students). People that didn’ t fullfil every step, didn’ t finish OSCE test or did not sign the informend consent form were excluded of the amount (21 students).

The amount was evalutated in three moments: two times via OSCE test with questions related about SPIKES protocol (the begenning and the end of 2019), and one time via online questionnaire (appendix 2) to understend and identify sociodemographic factors that could influence in the previous OSCE results (by the end of this online forms it had an video about breaking bad news Link avaible: https://youtu.be/DkLTzOlkVdo. OSCE test was taking inside the Universidade Positivo in a simulation room organized for this evalutaion.

All the authors of this article took a trainning by a Psychologist professor of Universidade Positivo and became capable to apply, read and register, by using a valid checklist (appendix 4) students’ performance. Four of authors of this article were together with the evaluator of each board of “breaking bad news”. In these stations, students were avalated by the authors of this article and the evaluators, whom had a valid checklist (appendix 4). There weren’ t interference between authors and evaluators’ evaluation.

All the sutdents had one minute to read the question on the door of the simulator room before go into the door, and had five minutes to realize the task. By the end of the simulation a sonorous signal indicated that the student had to change the station. This sequence was repited three times until all the students had passed to all the stations. After the end of the OSCE test, all participants had a debriefing, which is a discussion with the evaluators about their difficulties, mistakes, complications and impressions about the questions.

In the first OSCE test, the breaking bad news was a history of a 57-year-old man that had a gastric biopsy result indicated a high-level gastric adenocarcinoma. The student has to read the exam during the first minute of the station and then informed the pacient about this new.

In the second OSCE test, the breaking bad news was a history of a 32-year-old woman that had a BIHADS IV mammoraphy and a previous biopsy indicated breast cancer. The sutdent had to informed the patient about this new. Both of simulation had training actors played as patients.

A data bank was builed to verify all erros. It was used the StatisticalPackage for the Social Sciences (SPSS), 2.1 version software to statistics analysis of the data. All the statistics variables were descreptively analized. Numeric variables were calculated by using avarage and standard deviation. Qualitative variables were calculated by using relative and absolute frequencies. To verify equal hypotesis between groups’ avarage it was used the T test when distribution was normal, and Mann-Whitney test when normal test was refused. The normal test was the Kolmogorov-Smirnov. To prove the homogenicity of the groups acording to proportions, it was used Qui-square, or Fisher test to frequency above five.

## MATERIAL

For this study 12 rooms divided into four boards with three station each was used. The amount should read the question by using the limited time and should finished the task during the determine period. The results data collected, discussed and exposed only of medical students with regularized enrollment and participate of all three steps of the project (OSCE #1, OSCE #2, ans Online forms). Students whom missed one or more steps were excluded of the data.

The online questionnaire was written based in a breaking bad news and medical students article (ALBUQUERQUE, 2013). It was made through Google Docs tool (https://bit.ly/questionarioTCC).

This form was composed by 29 questions, divided into 26 questions that needed objective answers and three dissertative questions. It was divided into five parts: sociodemographics data; knowlegde of non-techenique and personal experience of them; personal expiriences related to breaking bad news; knowlegde of behavior in a breaking bad news situation; and knowlegde about SPIKES protocol, incluiding difficulties in use it.

Students had their performance registerd for each author of this article during the OSCE test execution, and the checklist data were written in a two-point form (0 = non success / 1 = success). Each item of the evaluation checklist was pondered, as indicated by the reference article, to calculate the students scores.

The total score used to evaluate the students was build based in SPIKES protocol. The score was divided into the itens that could worth 0.1, 0.5, or 1.0 points. The highest pontuations were attribuied to the items that were considered most important for the patients feelings about the news, and also items that help the patient to be positive despite the diagnosis.The addition of the scores in each iten was compared with the global score of all the OSCE’ s participants.

To analize these data, it was considered excelent efficiency up to 90% (scores > 4.5); good efficiency between 80% and 90% (scores between 4.0 and 4.4); moderate efficiency between 70% and 80% (scores between 3.5 and 3.9); and low efficiency under 70% (scores < 3,4). Every student that scored at least 3.0 points were considered up to the institutional mean (6.0).

## RESULTS

The 5th year of med school had 106 students with regularized enrollment. It was excluded of this amount students who didn’ t take the two OSCEs (9 students), who didn’ t answer online questionnaire (10 students), or those who had difficult with the sound system and couldn’ t be evaluated during the test (2 students). The amount for this article was 85 (n=85). Sampling error of 5% and confidence interval of 95%.

35 to 85 students were identified as males (44,7%), and 47 as females (55,3%). The age’ s median was of 25,19 years (minimal age of 22 and maximal age of 39 years).

About CMN, 50 students (58,8%) felt confortable with the breaking bad news situation; 73 students (85,9%) already experienced a CMN situation (avarege of 5,28 times +/-5,39): 50 of those considered the physician prepered to tell the patient the bad new; 60 of those reletaded that they had to talk to the patient about the new, resulting in an avarege of 1,97 times (+/-2,17). Up those 60 students 28 (32,9%) answerd that they felt prepred to this situation and 49 (87,5%) releted to follow an estrategy previous estabilished.

During the CMN, 65 students (76,4%) sad that the patient have to known everything related to her/his health; 79 (92,9%) sad that the doctor should know if the patient wants to know everything; 75 (88,2%) sad that the prognosis of the disease can influence the CMN; 80 (94,1%) sad that patient’ s perception of the disease is an important information for the doctor and 67 (78,8%) of the students sad that is normal the doctor feel emotion after the CMN. 79 students (92,9%) reported previous contact with SPIKES protocol, and the relation between the difficulty and facility with it.

The firts OSCE has 52% of studentes with results up to the institucional avarege. The highest score was 4,9 and the lowest score 0,5, an avarege of 3,02 with standard deviation of 1,05. The second OSCE had 59% of students up to the instititional avarege. The highest score was 4,7 and the lowest score was 0,9, an avarege of 2,94 and standard deviation of 1,09.

Relevant statistics diferences weren’ t observed between firts and second OSCE scores, as well as gender and age difference (under 25 or up to 25 years).

To compare the students’ difficulty level during OSCE exam about each item of SPIKES protocol, each item was divided into their referance in the protocol. A percentage score was calculated based on each checklist topic. The avarege and standard deviation of the first OSCE was: 76%(29) in Strategy, 80%(40) in Perception, 65%(27) in Knowledge, 67% (33) in Emotions, 28% (29) in Summary; the Qui-square comparion was relevent (P < 0,001). In the second OSCE, same data had the foowing percentage: 74%(29) in Strategy, 78%(41) in Perception, 67%(33) in Knowledge, 72% (32) in Emotions, 22% (23) in Summary; the hipotesis test had the same result in the first test when compered to the seconde one.

When analized over the previous expirience in CMN, medical students that already given the bad news had a pontuation avarege of 3,11 +/-0,84 in the second OSCE and those that releted never expirience CMN had a pontuation avarege of 2,57 +/-0,90. There wasn’ t statistic diference between gender (p = 0,346 in the first OSCE and p = 0,051 in the second one), between students under or up to 25 years (p=0,051 in the first OSCE and p=0,495 in the second OSCE) and between students that releted act following a plan already stablish and those who didn’ t (p=0,220 in the first OSCE and p=0,803 in the second OSCE). The differeces related are expressed in table 1.

**Table 1.** Quantity of right scores per criteria in both tests.

|  | First Test | Second Test | *P* |
| --- | --- | --- | --- |
| Q1. introduced himself to the patient | 64 (75,3%) | 73 (85,9%) | 0,081 |
| Q2. was interested in knowing the evolution of the disease since the last consultation | 63 (74,1%) | 69 (81,2%) | 0,269 |
| Q3. made a brief announcement of the problem | 68 (80,0%) | 52 (61,2%) | 0,007 |
| Q4. did active listening | 69 (81,2%) | 67 (78,8%) | 0,701 |
| Q5. reported the result properly | 55 (64,7%) | 43 (50,6%) | 0,063 |
| Q6. demonstrated tranquility when informing the result | 70 (82,4%) | 59 (69,4%) | 0,049 |
| Q7. made silence necessary for the patient's reaction | 60 (70,6%) | 39 (45,9%) | 0,001 |
| Q8. made a short explanation with the necessary information; possibility of therapy | 60 (70,6%) | 61 (71,8%) | 0,866 |
| Q9. made a short explanation with the necessary information; possibility of cure | 33 (38,8%) | 51 (60,0%) | 0,016 |
| Q10. performed active listening | 64 (75,3%) | 66 (77,6%) | 0,718 |
| Q11. reflected thoughts and paraphrased the words of the patient | 43 (50,6%) | 55 (64,7%) | 0,063 |
| Q12. offered comfort to the patient | 66 (77,6%) | 67 (78,8%) | 0,853 |
| Q13. not "gilded the pill" | 68 (80,0%) | 55 (64,7%) | 0,026 |
| Q14. did not avoid communication | 63 (74,1%) | 45 (52,9%) | 0,004 |
| Q15. did not "hang" himself | 60 (70,6%) | 56 (65,9%) | 0,510 |
| Q16. was not confusing | 50 (58,8%) | 47 (55,3%) | 0,642 |
| Q17. did not interrupt the patient | 66 (77,6%) | 67 (78,8%) | 0,853 |
| Q18. kept the patient's hopes up | 51 (50,0%) | 65 (76,5%) | 0,021 |
| Q19. was empathetic to the patient | 55 (64,7%) | 61 (71,9%) | 0,323 |
| Q20. asked if the patient had any questions | 36 (42,4%) | 29 (34,1%) | 0,269 |
| Q21. made himself available for further queries | 31 (36,5%) | 23 (27,1%) | 0,188 |

Qui-square, Pearson and Fisher Test were used in this article, as well as T Test to equality of means and ROC curve. P values under 0,05 were considered significant

## DISCUSSION

To achieve the highest quality of medical education, methods and activities about CMN are required. The SPIKES protocol is one of the tools that is easy to use and access, and makes the CMN learning less confused (FERREIRA DA SILVEIRA; BOTELHO; VALADÃO, 2017; LINO et al., 2011). Theory classes about CMN are already part of the curriculum, but it is not enough (COUTINHO; RAMESSUR, 2016).

A systematic review showed that blanded methodos, which use practice and theory to teach has bigger importance to teach CMN (CAMARGO et al., 2019). The active teaching between PS in the OSCE test, for example is an promising methods to evaluate and permits that students have a final feedback to improve their learning (DOURADO; GIANNELLA, 2014; HARDEN et al., 1975; ROSENBAUM; FERGUSON; LOBAS, 2004; SCHWELLER et al., 2018). The actual article realized experiences with PS trained to act accordingly with the literature, exposing their feedback and make them comfortable to make questions (FORTIN et al., 2002).

This study demonstrated that the students achieved a medium performance in the knowledgment of breaking bad news. Both of OSCEs a little more than half of the students achieved scores above the institutional average, 52%(n= 44) in the first OSCE and 59% (n= 50) in the second OSCE. The general average also reports medium performance, and also has reduction between the two tests (from 3,02 (dp= 1,05) to 2,94 (dp= 1,09)) with no statistical significant (p=0.0809). It wasn’ t possible to evaluate significant enhancement between OSCEs average. This was an expected result in this study. Other similar articles that use this type of evaluation of CMN experience better performance between the tests. Setubal et al(SETUBAL et al., 2018), separated, in his study perinatology residents into control and intervention groups. The intervention group received a 1-2h30 hour of SPIKES protocol training. It was verified that there was not a significant difference between groups’ performance. In the second simulation both groups got better performances, though. That shows that previous practice and experiences enhanced their performances (SETUBAL et al., 2018).

By the end of the simulations, 58,8% (n = 50) of the students were considered comfortable with breaking bad news situations. There is an association between patients’ simulations training and confidence development in the moment to use CMN in real situations (LIFCHEZ; REDETT, 2014). Lifchez et al used in his study plastic surgery residents and identified that their confidence, as well as their performance enhanced after PS training. This fact was also observed by the test evaluators. By the end of this study, it was concluded that this type of simulation is very important (LIFCHEZ; REDETT, 2014). Karam et al identified in his study an increase (56,2 to 93,7%) in the “good” scores in breaking bad news after the practice workshop (KARAM et al., 2017). Similar thing probably is going to happen with this study, because by the end of the graduation all the students are going to participate in three other OSCEs.

The online questionnaire that was used was also applied, by another study, to medical students of every year of graduations of multiples universities of Portugal (ALBUQUERQUE, 2013). In this present study 85,9% of patients already had experienced a breaking bad notice situation, while in the Portuguese study about 75% already had the same experience. In the Albuquerque article the most difficult item of SPIKES protocol, according to the med students was “Emotions”, the same item reported in this study. Although our students reported this difficulty, this was not compatible with the scores in the OSCE test. When compared with the other SPIKES protocols, “Emotions” was the third item that students got the highest score in the two OSCEs. Armin Azadi had a similar finding, dealing with the patient’ s emotions was reported as the hardest part of BBN where in fact it got the highest score of the SPIKES items (Azadi et al.,2019).

When comparing the checklist items, it is noticed that there was a decrease in the number of correct answers in question number six (was the student calm when informing the results to the patient?). In the first simulation there were 76 (82,4%) correct answers, while in the second simulation there were only 56 (69,4%) correct answers (p=0,049). One of the reasons that this happened is that in the second simulation the student received the exam result inside the room. The same reason can also justify the decrease (16) in the number of correct answers in question number three (did the student make a short announcement?) (p=0,07).

These differences between the evaluation rooms lead to a higher level of difficulty in the second OSCE, and this fact justifies the higher number of wrong answers in questions number 7, 13, 14, and 25.

Questions related to the end of BBN had the least numbers of correct answers in both OSCEs (questions number 20, 21, 22, 23, and 24). This fact points that the time settled to this question-five minutes - was insufficient. In the first OSCE question number 23 has the highest number of incorrect answers; in the second OSCE the same question has only nine correct answers. In Setubal study (SETUBAL et al., 2018) residents needed 12 minutes to finish this task.

When comparing these results to a previous study that used the same checklist, there are a few differences. The first thing to notice is that the most number of wrong answers were related to not encouraging the patient despite the bad news (SOMBRA NETO et al., 2017), which is different from this study. Another important difference is that there is an important difference between the performances: 67% (n= 80) of students had excellent performance in Sombra Neto study (SOMBRA NETO et al., 2017) and in this present study there are moderate performances. This fact can be explained because of method differences.

In Sombra Neto study (SOMBRA NETO et al., 2017) students had to go to weekly theoretical classes and simulations classes about breaking bad news for about one month. Students knew which type of patient they had to talk about the disease. This intervention did not occur in the present study. Because of that we have a big difference in scores and performances.

Gender and age are not elements that influence scores. There are no studies analyzing these two elements as important factors to increase scores in performance.

Also there is no statistical significance between using a protocol and getting high scores. Carvalho Caleffi relates that every breaking bad news situation has to be analyzed as a single situation and sometimes protocols are not appropriate for BBN (CARVALHO CALEFFI et al., 2016). The present study agrees with Carvalho Caleffi because there is no statistical significance when students used something planned to communicate bad news.

The previous experience was a factor that showed to be statistically significant for the second OSCE score. This seems to be related to an increase in the performances. Some studies evidence that CMN training and previous experience are important to improve this skill. Arnold (ARNOLD et al., 2015) identified that residents felt more confident in CMN and their performance increased up to 23% after practice training. Colletti (COLLETTI et al., 2001) showed in his study that students who received previous training had the highest scores. It is recognized that the present study has limitations. The difficulty variation in OSCEs, as well as the 6 month gap between them contributed to make conclusions about scores evolution and performances. The initial project included at least three OSCEs with breaking bad news questions but just two of them were realized due to COVID-19 pandemic.

The debriefing time by the end of the exam was around 15 minutes. This time can be considered short because at this moment all OSCEs questions were discussed with students. Other studies reported that it is necessary between 30 and 40 minutes to this discussion (SKYE et al., 2014). There is only one Med School involved in this project, and because of that there is a quality limitation. Despite these limitations the present study is one of the first studies using these checklists in OSCEs evaluation, as well as being one of the first studies to analyze the importance of training this unique skill, specially in a pandemic year where there are unfortunately a lot of bad news.

## CONCLUSION

Unfortunately, it was not possible to compare students’ performance in the first and the second test due to previous discussed test limitations. It is necessary to investigate if the method contributes to these limitations.

Although the majority of students know SPIKES protocol there are no statistical significance in the previous acknowledgement of the protocol and high scores in tests.

The only factor that positively influenced the student’ s performance was the CMN previous experience.

The study revealed OSCEs potential as an education tool. Material and methods of this study indicate that this type of study can be developed with similar methods.

## Data Availability

All data produced in the present work are contained in the manuscript.

## ABREVIATIONS’ LIST

OSCE: *Objetive Estructured Clinical Examination*
CMNBreaking: bad news
PSSimulators: patients

